# Validation of automated hippocampus volume assessment using deep learning convolutional neural networks in patients with Alzheimer’s disease

**DOI:** 10.1101/2023.04.11.23288432

**Authors:** Nur Shahidatul Nabila Ibrahim, Subapriya Suppiah, Buhari Ibrahim, Nur Hafizah Mohad Azmi, Vengkatha Priya Seriramulu, Mazlyfarina Mohamad, Marsyita Hanafi, Hakimah Mohammad Sallehuddin, Rizah Mazzuin Razali, Noor Harzana Harrun

## Abstract

**Background:** Dementia is a spectrum of diseases characterised by a progressive and irreversible decline in cognitive function. Appropriate tools and references are essential for evaluating individuals’ cognitive levels, especially hippocampal volume as it is the commonly used biomarker in detecting Alzheimer’s disease (AD). It is important to note that while there is no cure for dementia, early intervention and support can greatly improve the lives of those affected.

**Method:** Ongoing research is being conducted to develop new treatments and improve our understanding of the disease by using VBM to compare sensitivity and specificity with the HippoDeep toolbox.

**Result:** We were able to validate AD’s hippocampal volume compared to age-matched healthy controls (HC) based on HippoDeep Model by comparing it with VBM as the reference standard.

**Conclusion:** There are significant differences between hippocampal volume in AD and HC that have been detected using VBM and HippoDeep analysis.

## 1.0 INTRODUCTION

AD is the commonest neurodegenerative disorder that affects older adults and leads to significant morbidity. A combination of clinical assessment, neuropsychological tests, and diagnostic imaging to help increase diagnostic accuracy. One of the hallmarks of AD is hippocampal atrophy which is detected using magnetic resonance imaging (MRI). Nevertheless, the measurement method is numerous and can give rise to variable results.

Historically, the hippocampal volume measurement was made using 2D slices on MRI images acquired on a high-resolution T1-weighted MRI sequence. Eventually, this progressed into developing the hand-drawn volume of interest (VOI), which segmented the brain and selected the hippocampus for volumetric assessment. Several image processing software has also been designed to evaluate hippocampal volume; for example, the Statistical Parametric Mapping (SPM) tool can be used to process raw structural data to measure the volume of the hippocampus. SPM can be used to assess Voxel-based Morphometry (VBM) of the hippocampus in a semi-automated manner. These techniques have inherent limitations as they are time-consuming, tedious to perform and require a certain amount of technical training. The measurements are potentially inconsistent and depend on the person performing the measurements. Thus, artificial intelligence may be the solution to provide more consistent, accurate, and reproducible results. Using deep learning, the convolutional neural network (CNN) algorithm permits an automated pipeline to be run and measure more data in a shorter period. One such CNN-based algorithm is the HippoDeep model(x). This model was designed to perform automated segmentation and measurement of hippocampal volume.

Our study’s objective was to validate AD’s hippocampal volume compared to age-matched healthy controls (HC) based on HippoDeep Model by comparing it with VBM as the reference standard. We hypothesise that HippoDeep Model had a good correlation and comparable diagnostic accuracy with the VBM method.

## 2.0 MATERIALS AND METHODS

### Subjects’ recruitment and setting

This is a cross-sectional case-control study that was conducted from February 2021 to March 2023. The AD subjects were recruited from the memory clinics at the Geriatric Clinic in Hospital Kuala Lumpur, Klinik Kesihatan Pandamaran in Klang, and Hospital Sultan Abdul Aziz Shah Universiti Putra Malaysia. These AD subjects were diagnosed based on the clinicians’ examination and neuropsychological testing using various cognitive assessment instruments, such as the Mini-Mental State Examination (MMSE) and the Montréal Cognitive Assessment (MoCA) questionnaires, overall being guided by the DSM-5 criteria for major neurodegenerative (First et. al., 1975). The cognitively healthy HC subjects were recruited through advertisements in local bulletin boards and flyers distributed to the community.

### Sample size determination

The calculated sample size was based on the data presented by Buhari et al., 2021. Whereby, the prevalence of AD in this region was 25%. Using Arifin, W.N.,2023, we utilised a web-based sample size calculator downloaded from http://wnarifin.github.io., giving a minimum sample size of 30 subjects per group.

### Ethical clearance

This study was approved by the Medical Research Ethics Committee (MREC) of the National Medical Registration Registry (NMRR) Malaysia (NMRR-19-2719-49105) and the Ethics Committee for Research Involving Human Subjects of Universiti Putra Malaysia (JKEUPM-2019-328).

### Neuropsychological testing

All the recruited subjects underwent our trained researchers’ MMSE, MoCA and CDR assessment. The values were interpreted as MoCA: >26 as cognitively healthy; 16.3-25 as moderate cognitive impairment; <16.2 as having severe cognitive impairment, MMSE: 24-30 as cognitively normal; 18-23: moderate cognitive impairment and 0-17: severe cognitive impairment: As for the CDR that evaluated the effect on the activities of daily living, the assessment was based on a questionnaire with, No Dementia (CDR = 0), Questionable Dementia (CDR = 0.5), MCI (CDR = 1), Moderate Cognitive Impairment (CDR = 2), and Severe Cognitive Impairment (CDR = 3).

### Protocol for structural MRI imaging

High resolution, T1-weighted MRI scans were performed using a 3.0-T SIEMENS Prisma scanner with software version Syngo MR D13D, having TR: 2300 ms; TE: 2.27 ms; Inversion time: 900 ms; Slice thickness: 1 mm; Number of slices: 160; Flip Angle: 8.

### Pre-processing using SPM Matlab

All the T1-weighted images were preprocessed using SPM Toolbox (https://www.fil.ion.ucl.ac.uk/spm/download/restricted/eldorado/spm12.zip) implemented in MATLAB (R2021a).

### Processing VBM

The steps were adopted from the (VBM protocol), beginning with segmentation (spatial pre-processing): using the native space option, the researcher can generate a tissue class image aligned with the original. Next, execute Dartel to generate templates from selected images to be distorted. The next step involves normalisation, where the tissue class image is transformed into a standard space, followed by smoothing to improve the signal-to-noise ratio. These steps are crucial in preparing the data for further analysis. Images with the same dimensions, orientation, and voxel size were selected in order to perform statistical analysis using the factorial design specification. Using a design specification file from the computer’s file system, the cost was calculated. The Automated Anatomical Labeling (AAL) toolbox was superimposed with the Montreal Neurological Institute (MNI) template to extract the volume to cluster level.

### Processing for HippoDeep using FSL

HippoDeep relies on a hippocampal appearance model learned from existing FreeSurfer v5.3 labelled online data sets and synthetic data rather than warping individual images to an atlas (Thyreau et al., 2018). The HippoDeep CNN is trained using two different kinds of synthetic data. The first artificial data set consists of a manual segmentation of a synthetic high-resolution image of the hippocampus created from an average of 35 consecutive MRI scans of a single healthy participant. The goal of segmenting the hippocampus on a high-resolution image (0.6 mm isotropic resolution) was to give the CNN more detailed boundary information that may need to be clarified on a lower-resolution image. The second type of synthetic data used to train the HippoDeep CNN is FreeSurfer v5.3 training data that has been artificially geometrically warped. While some of the distortions were outside the range of clinically acceptable values, it is still realistic enough for a human rater to be able to distinguish them. This distorted data was used to give the CNN practical training recommendations. Details on the specifics of how the synthetic data were generated may be found in the study by Thyreau et al. (2018).

HippoDeep Brain hippocampus segmentation installation were done based on using a Repo for ubuntu/Linux and Mac OS compatible:https://github.com/bthyreau/hippodeep_pytorch. Whereas, for a Windows-based system, the repo is https://github.com/bfoe/hippodeep_pytorch. We utilised executable Windows pre-compiled app: https://github.com/bfoe/hippodeep_pytorch/releases/download/v0.3/HippoDeep_Windows_v0.3.zip It is a plug-and-play pre-compiled app for use on Windows platform.

HippoDeep_Windows_v0.3.zip

Once we installed the HippoDeep toolbox, we opened the high-resolution T1-weighted images and followed the instructions in the pop-up box. The processed images and reports were saved in the original T1-weighted file.

### Statistical analysis

The statistical tests were conducted using Statistical Package for the Social Sciences (SPSS software Version 23.0, SPSS Inc., Chicago, IL, USA) and MedCalc, a statistical software package designed for the biomedical sciences. It has an integrated spreadsheet for data input and can import files in several formats. The level of significance was set at p-value < 0.05. Spearman correlation was used to determine the association between the VBM and HippoDeep measurements of bilateral hippocampi volumes of AD and HC subjects. ANCOVA test was used to analyse the association between the hippocampi volume and age. The ROC curve was used in this study to compare the accuracy of hippocampus volume measurements between VBM and HippoDeep methods.

## 3.0 RESULTS

The final data was lower than our calculated sample size because we selected the best-suited age-matched subjects for this evaluation. Table 1 shows the age and neuropsychological test scores distribution based on the mean values and standard deviation between AD and HC groups.

**Table 1.** Distribution data for HC and AD

|  | HC | AD |
| --- | --- | --- |
| n | 15 | 15 |
| Age | 71.2 (7.68) | 74.27 (10.05) |
| MoCA | 27.33 (3.13) | 14.13 (6.82) |
| MMSE | 27.53 (2.61) | 14.4 98.33) |
| CDR | 0 90.00) | 2.07 (0.88) |

**Table 2.** Bilateral hippocampal volume difference in VBM and HippoDeep.

| Variable | Mean (SD) |  |
| --- | --- | --- |
|  | VBM | HippoDeep |
| Left Hippocampus Volume | 1.39 (0.43) | 2.55 (0.60) |
| Right Hippocampus Volume | 1.47 (0.38) | 2.68 (0.66) |

Spearman rank correlation was computed to assess the association of hippocampus volume using VBM method and HippoDeep Toolbox. There was a positive strong correlation between the two variable, r (28) = 0.612, p < 0.001 using VBM method, while there was also a positive very strong correlation between the two variable, r(28) = 0.937, p < 0.001 using the HippoDeep method.

A One-way ANCOVA was conducted to determine whether there was any significant difference between the hippocampal volume of the AD and HC based on VBM and HippoDeep methods using age as a covariate. There was a significant difference between the left hippocampal volume between the AD and HC group using VBM analysis, F (1, 29) = 0.942, p< 0.001. There was also a significant difference in left hippocampal of AD and HC using HippoDeep toolbox, F (1, 29) = 1.08, p< 0.001. One-way ANCOVA also detected a statistically significant difference between right hippocampal volumes in HC and AD groups using the VBM analysis and HippoDeep method. There was a significant difference between the right hippocampus volume of AD compared to HC using VBM analysis, F (1, 29) = 0.795, p < 0.001. While using the HippoDeep toolbox, there was also significant difference detected between the right hippocampal volume of the AD compared to HC groups, F (1, 29) = 1.403, p < 0.001.

Figure 1 and Figure 2 show the scatter plot pairs of numerical data of hippocampal volume distribution according to age for the VBM and HippoDeep-based methods, comparing the AD and HC groups distribution to demonstrate the relationship between them. The linear line shows the pattern of a decrease in hippocampus volume with increasing age, as this variable is inversely proportional to age. Furthermore, the values of right and left hippocampal volume, measured using both these methods, showed that these values were lower (showing more accelerated atrophy) in the AD group compared to the age-matched HC group.

**Figure 1.**
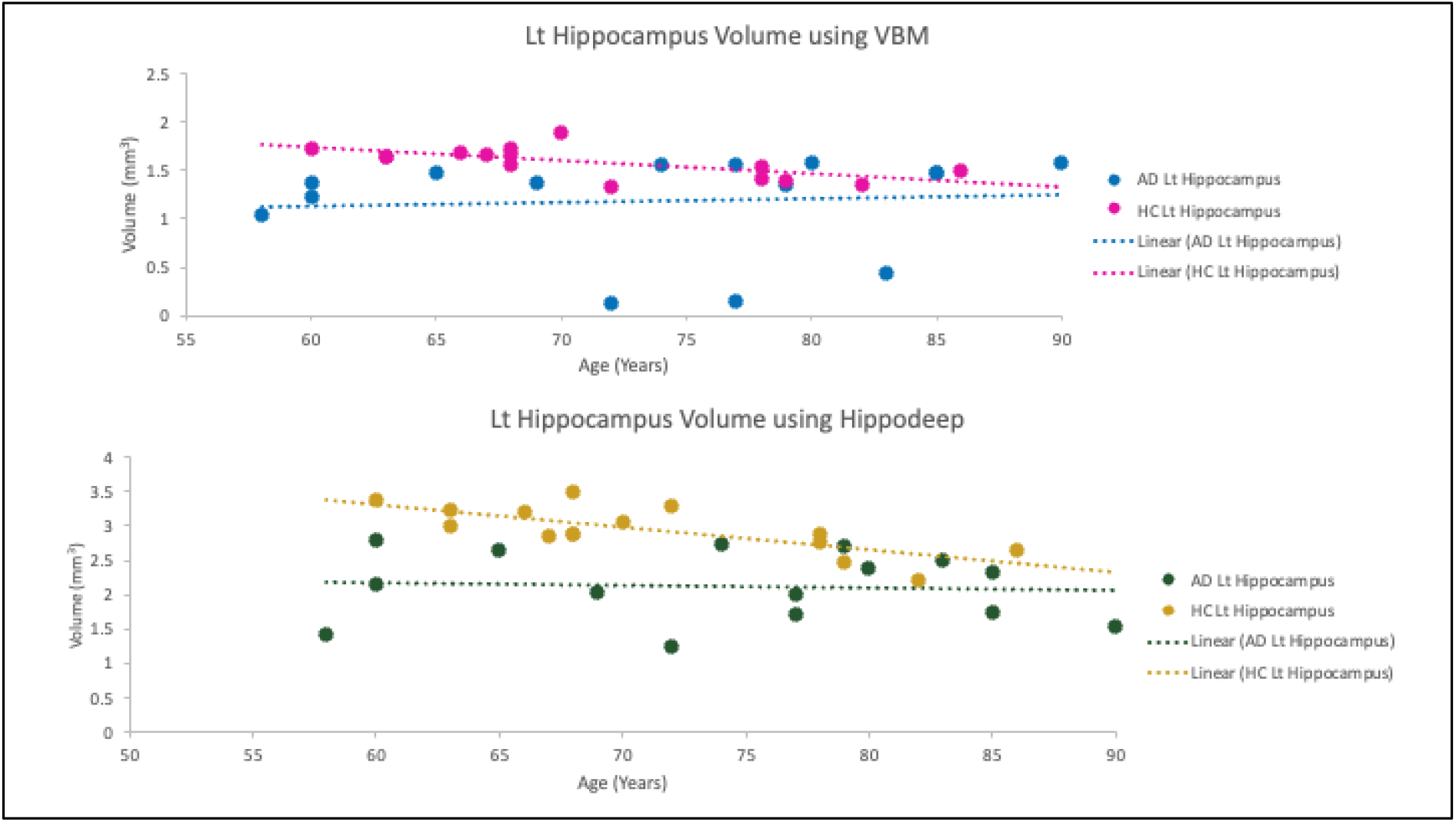
Left hippocampal volume distribution according to age using VBM and HippoDeep analysis for AD compared to HC groups.

**Figure 2.**
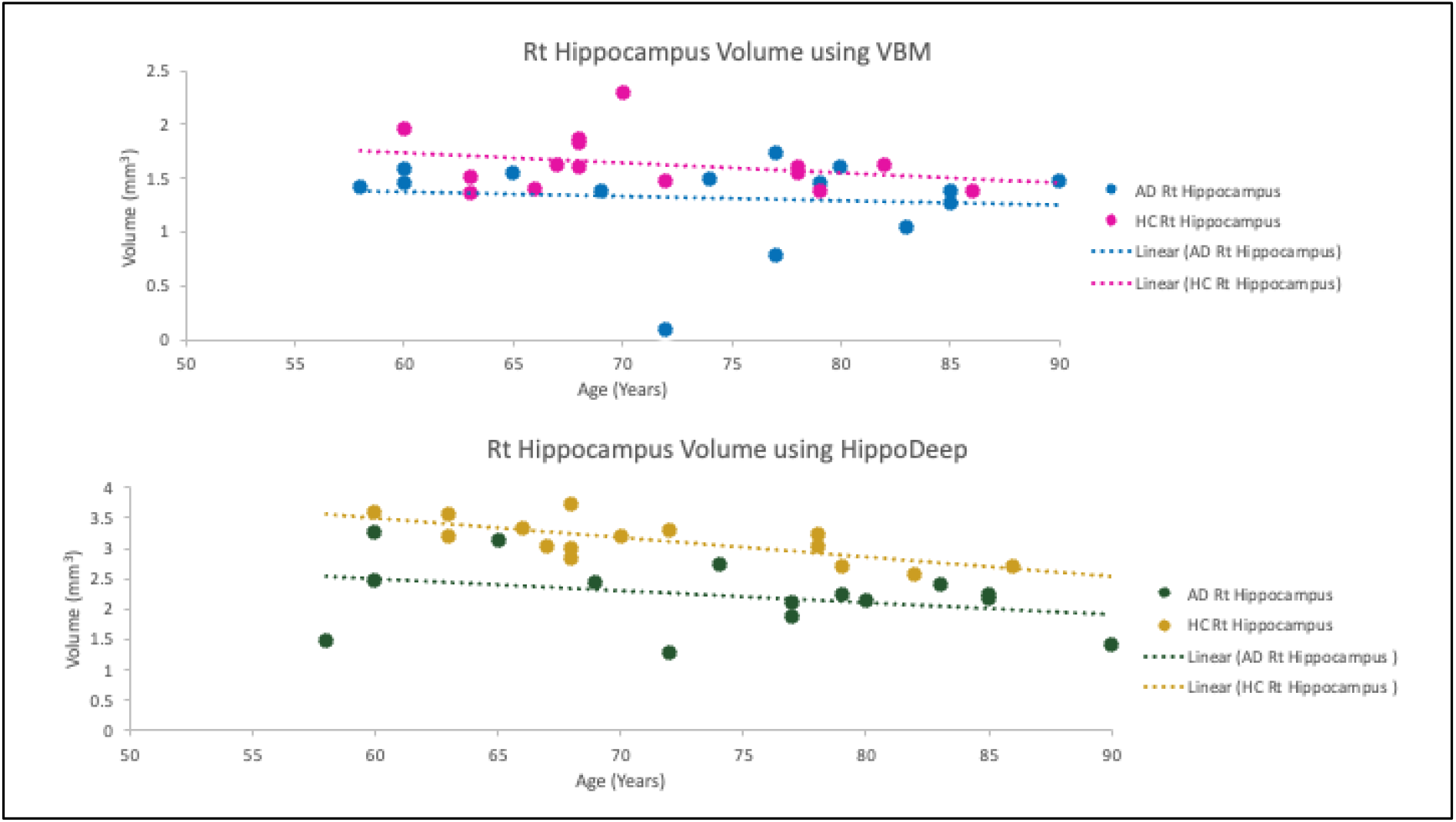
Right hippocampal volume distribution according to age using VBM and HippoDeep analysis for AD compared to HC groups

**Figure 3.**
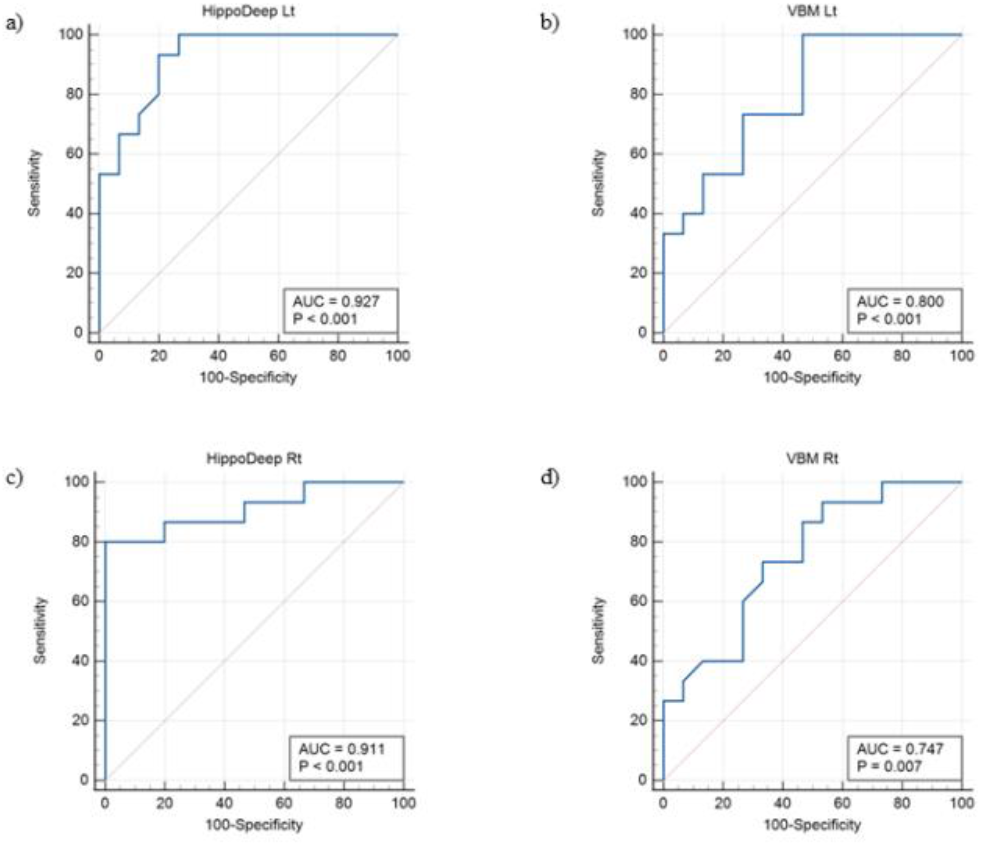
ROC curve of the accuracy of the HippoDeep compared to the VBM model for classifying Alzheimer’s disease based on the measurements of bilateral hippocampal volumes. (a)HippoDeep Left Hippocampus Volume, (b) VBM Left Hippocampus Volume, (c) HippoDeep Right Hippocampus Volume and (d)VBM Right Hippocampus Volume

## 4.0 DISCUSSION

Individuals with dementia are assessed for cognitive dysfunction using the Mini-Mental State Examination (MMSE) and the Montreal Cognitive Assessment (MoCA). Clinicians and researchers require norms for cognitive tests in order to assess the cognitive levels of subjects referred for evaluation (Riello et. al., 2022). MMSE and MoCA are significantly different in AD and HC based on the study in 2022 by Wei et. al. Show the measurement have done data pooled with community-based shows the different cognitive levels between cognitively healthy and dementia patients.

Fundamentally, the most salient structural change that is ubiquitously present in AD patients is the atrophy of the hippocampus in the medial temporal lobe. The hippocampal volume in AD is significantly smaller in AD compared to HC even after being corrected for increasing age. In a study by Hu et al. (2023), it was mentioned that MRI-estimated hippocampal volume is a superior structural biomarker than total medical temporal lobe volume or entorhinal cortex volume. The European Federation of the Neurological Societies (EFNS) (Hort et al., 2010), the EMA (Hill et al., 2014), the National Institute on Aging and the Alzheimer’s Association (NIAAA) (Albert et al., 2011), and the International Working Group (IWG) (Dubois et al., 2014) recommended hippocampal volume as a supplementary biomarker facilitating the clinical diagnosis of AD. HippoDeep can differentiate the two groups based on hippocampal volume in a simplified and automated manner.

HippoDeep was able to predict and classify AD compared to HC better as evidenced by the ROC curve of left hippocampal volume with sensitivity 93.33. % and specificity of 80.00% and AUC 0.927 while right hippocampal volume with sensitivity 80.00%, specificity of 100.00% and AUC 0.91. This is in comparison with VBM methods that gave the best balance of left hippocampal volume sensitivity of 100.00%, specificity of 53.33% and AUC 0.800 while in right hippocampal volume sensitivity of 73.33%, specificity of 66.67 and AUC 0.747. Another study in 2016 done by DeMarshall et al. proved that ROC is able to detect disease specificity of the selected biomarkers for AD using ROC curve assessment. In a study by Suzuki et al. (2023) it was notedthat area under the curve (AUC) was more than 0.8, indicating a good sensitivity and specificity in general for classifying AD. HippoDeep can better predict and classify AD compared to HC by measuring the right hippocampus volume with an accuracy of 91.1%, compared to VBM, which has 74.4% accuracy for the same side of hippocampal volume.

Interestingly, our study also detected that the left hippocampus demonstrated a more significant decrease in volume compared to the right, which is the magnitude of non-directional hippocampal asymmetry that the decreasing cognitive state may accelerate. Moreover, *in vivo* MRI volumetry research consistently showed that the right hippocampus is larger than the left in large population studies (Pedraza et al., 2004)

## 5.0 CONCLUSION

We validated the HippoDeep tool as an alternative method to VBM analysis for measuring the hippocampal volume. HippoDeep has a higher sensitivity and specificity than the VBM analysis method, enabling it to accurately classify Alzheimer’s patients while consuming less time and requiring minimal training.

## Data Availability

All data produced in the present study are available upon reasonable request to the authors

## Acknowledgements

This research was funded by the Ministry of Higher Education, Malaysia under the Fundamental Research Grant Scheme (FRGS/1/2019/SKK03/UPM/02/4) and project code: 04-01-19-2119FR and project number 5540244 that was awarded to Associate Professor Dr. Subapriya Suppiah (Geran Kementerian Pengajian Tinggi, Malaysia, nombor kod rujukan FRGS/1/2019/SKK03/UPM/02/4 dengan nombor projek 5540244)

## Author Contributions

SS conceptualised the study design. NSNI carried out the literature search, data extraction, and quality assessment. BI, NHMA and VPS carried out the data collection. NSNI wrote the manuscript’s first draft. SS, MH, MM, and HMS edited the manuscript, verified the data, and provided critical feedback to help shape the research. RMR and NHH are clinicians who refer their patients as research subjects in this study.

